# Mapping Stakeholder Alignment for Deprescribing Policy in France: Insights from a Policy Delphi Approach

**DOI:** 10.64898/2026.02.11.26346080

**Authors:** Sergio Oliveira, Odessa Dariel, Matthias Brunn

## Abstract

**Background:** With growing efforts aimed at optimizing health care services by reducing “low value care”, medical deprescribing represents a critical policy challenge at the intersection of clinical quality, fiscal sustainability, and environmental stewardship. Despite growing evidence of its benefits, France lacks a comprehensive national framework for systematic medication review and deprescribing implementation.

**Objective:** To identify areas of consensus and divergence among key French stakeholders using an adapted Policy Delphi approach to inform national deprescribing policy development.

**Methods:** An exploratory survey was conducted among stakeholders across five groups (healthcare professionals, patients, academia, policymakers, and the pharmaceutical industry). Consensus levels were assessed using graded Likert scales and analysed across policy domains, including knowledge and training, collaboration, resources, policy support, and sustainability opportunities.

**Results:** High consensus emerged around knowledge gaps, the need for interprofessional collaboration, and clinical benefits of deprescribing. Moderate consensus existed regarding resource constraints and environmental sustainability. Divergence was observed between professionals/academia and policymakers/industry regarding financial incentives and regulatory readiness. A policy Delphi heatmap revealed specific alignment patterns that could serve as policy entry points.

**Conclusions:** Multi-stakeholder consensus mapping provides an innovative governance tool for identifying actionable policy opportunities and contributes to recent tools aimed at reducing low-value care. High-consensus domains, including training, patient safety, and sustainability, offer immediate entry points for coalition-building. On the contrary, areas of divergence require structured dialogue and iterative policy learning among France’s fragmented governance structures to translate stakeholder alignment into systematic deprescribing implementation.

## 1. Introduction

Healthcare systems worldwide face enormous pressures from population ageing, rising chronic disease prevalence, and escalating pharmaceutical expenditures. Yet recent estimates suggest that ‘low-value care^1^’ accounts for an increasingly large portion of healthcare spending in high-income countries (Fraser et al., 2024; Warkentin et al., 2026). In France specifically, with a population over 65 approximating 22% and those living with multiple chronic conditions representing 46% of the population, only 20% of clinical consultations are prescription-free (INSEE, 2025; OECD, 2023; Richard et al., 2023). This results in frequent cases of polypharmacy and inappropriate medication use, affecting approximately 43% of older adults (Drusch et al., 2023). Polypharmacy is associated with a 20-30% increase in adverse drug reactions, leading to preventable hospitalisations that incur significant economic costs, with a paradoxical need for additional pharmaceutical spending that, overall, reached €33 billion in 2023 (DRESS, n.d.; Drusch et al., 2023; Laroche et al., 2017).

Among the different strategies aimed at reducing low-value care and increasing health system performance, deprescribing has emerged as a safe clinical intervention that has been associated with not only improved health outcomes and improved quality of life for patients, but also decreased healthcare utilisation (Carollo et al., 2024; Reeve et al., 2015, 2017). Furthermore, deprescribing supports environmental sustainability by reducing pharmaceutical waste and carbon emissions. Yet, despite this evidence, deprescribing remains a marginal practice in France as the country lacks a comprehensive national framework safeguarding these practices, with efforts fragmented across regional health agencies, insurance, regulatory agencies and professional organisations (12. Ministère des Solidarités et de la Santé, 2023; Daunt et al., 2023; Gouvernement de France, 2023; OMEDIT Pays de la Loire, n.d.; The Shift Project, 2023).

### The Policy and Governance Challenge

Understanding why deprescribing has not gained traction in France requires moving beyond clinical considerations to examine the policy ecosystem in which medication management decisions are embedded. France’s healthcare system is characterised by strong state stewardship combined with pluralistic provision, with governance distributed across multiple actors. The *Ministère du Travail, de la Santé, des Solidarités et des Familles* sets national strategy via the *Direction Générale de la Santé* (DGS), the *Haute Autorité de Santé* (HAS) evaluates medical technologies and issues clinical guidelines, the *Caisse Nationale d’Assurance Maladie* (CNAM) acts as policy enforcer and reimbursor and regional agencies implement policy at the local level (Brunn & Hassenteufel, 2021; Zeynep Or et al., 2023; Zeynep Or & Coralie Gandré, 2021). This fragmentation creates coordination challenges and implementation gaps, misaligning the professional, institutional and political spheres (Ailabouni et al., 2022; Simonet, 2021). Moreover, powerful interest groups with competing priorities shape the French healthcare policy ecosystem. Professional unions representing clinicians and pharmacists wield considerable influence and use it to advocate for clinical autonomy (Bras, 2008; François Buton, 2024); while patient advocacy groups promoting safety and patient rights run up against strong pharmaceutical lobbying for more favourable policies that often conflict with patient interests and deprescribing agendas (Khazzaka, 2019). Understanding the attitudes, priorities, and areas of alignment among these different stakeholders is essential to provide policymakers with the information they need to develop evidence-based policies to tackle low-value care through de-implementation strategies, such as deprescribing (Warkentin et al., 2026).

### The Evidence Gap

While deprescribing is still “niche” in research, literature has grown substantially in recent years, mostly focused on identifying common barriers and facilitators for deprescribing across the healthcare ecosystem (Charbonneau et al., 2024; Doherty et al., 2020; Gillespie et al., 2018). However, from a policy perspective, research is limited mainly to prescriber and patient behaviours and specific clinical interventions, leaving behind the analysis of the problem as a multi-stakeholder policy challenge (Heinrich et al., 2022; Okeowo et al., 2023). Notable exceptions are recent work by (Riordan et al., 2016) on stakeholder perspectives in Ireland and the Canadian Deprescribing Network’s consultation processes, but comparative policy analyses remain scarce (Barbara Farrell et al., 2017). Existing research rarely focuses on the perspectives of policymakers and industry actors, despite their critical role in shaping the pharmaceutical policy, including the EU Pharmaceutical Package or the Critical Medicines Act (Cross et al., 2021; Silva Almodóvar et al., 2024). Studies tend to report descriptive findings about stakeholder views rather than analysing patterns of consensus and divergence that could inform coalition-building and policy design (Bolt et al., 2023; Forest et al., 2021).

This study addresses these gaps by employing an adapted Policy Delphi approach to map areas of consensus and divergence among key French stakeholders on deprescribing. The idea is to build a “policy readiness map” visualising where stakeholder alignment enables action and where sustained dialogue and negotiations are required. By doing so, we hope to propose concrete *policy entry points* (Kanger et al., 2020) for advancing a national deprescribing framework in France. To date, this study provides the first comprehensive mapping of French stakeholder perspectives on deprescribing and a consensus mapping that can serve as a governance tool for policy development in complex, multi-actor healthcare ecosystems.

## 2. Methods

### Study Design and Theoretical Framework

We conducted an exploratory cross-sectional survey and an adapted Policy Delphi methodology to assess stakeholder perspectives on deprescribing in France. We performed a single-round survey with graded Likert-scale responses that allowed us to quantify levels of agreement within and across stakeholder groups. The survey was complemented by open-ended questions to capture nuanced perspectives and unexpected insights.

### Stakeholder Selection and Sampling

We identified five key stakeholder categories based on their capacity to influence deprescribing policy: (1) healthcare professionals, (2) patients and caregivers, (3) academia and research institutions, (4) policymakers and (5) pharmaceutical industry representatives. We conducted a comprehensive stakeholder mapping in March 2025, informed by literature on French health and pharmaceutical policy governance (Brunn & Hassenteufel, 2021). Stakeholders were targeted at national, regional and local levels to capture implementation perspectives across the country.

### Survey Development

We developed the survey through an informed multi-step process and expert consultation. We conducted searches in PubMed and Scopus for articles published from 2010 onwards using terms related to deprescribing, polypharmacy, stakeholder perspectives, barriers, and facilitators. This web-based research yielded 45 key articles including themes such as knowledge and training, clinical confidence and attitudes, patient perspectives, communication challenges, interprofessional collaboration, resource constraints, regulations, incentives, and both economic and environmental considerations. The questions were tailored to each stakeholder group while maintaining sufficient overlap to enable cross-stakeholder comparison. The survey consisted primarily of closed-ended questions using five-point Likert scales (1=Completely disagree, 2=Disagree, 3=Neutral, 4=Agree, 5=Completely agree) or binary yes/no responses for factual questions. A panel of key informants with expertise in the topic who were not eligible participants in the main study assessed the validity of the content.

### Data Collection

The survey took place from April 1 to May 19, 2025 and we contacted participants directly via email. To broaden the reach beyond the initial mapping, we designed a supplementary recruitment strategy employing LinkedIn posts inviting interested organisations to contact the research team. Upon confirming eligibility, these additional respondents received the survey link via email. Respondents provided organisational affiliation and stakeholder category identification. When multiple responses were received from the same national organisation, one was randomly selected for inclusion. For regional or local entities, all responses were retained, recognising that implementation perspectives may vary across French territories.

### Data Analysis

We used descriptive statistics for each stakeholder group to analyse response rates, distributions across Likert categories, and proportions for binary questions. We assessed consensus levels using an adapted Policy Delphi framework (Pauline Van Ngoc, 2025). Consensus was defined as high for ≥70% agreement in a single Likert category (agree or disagree) OR ≥80% agreement in grouped categories (agree + completely agree, or disagree + completely disagree), moderate for ≥60% and <70% agreement in a single category OR ≥70% and <80% in grouped categories, low for >50% and <60% in a single category OR ≥60% and <70% in grouped categories, and no consensus for all other distributions. We conducted a cross-stakeholder analysis by grouping questions into thematic policy domains (knowledge and training, feelings and values, resistance and barriers, communication and collaboration, resources, policy and regulatory preparedness and opportunities). For each domain, we compared consensus patterns across stakeholder groups to identify convergence and divergence.

### Ethical Considerations

According to the French Public Health Code and CNRS COMETS guidance, non-interventional social science research involving organisational and policy stakeholders does not qualify as “research involving humans” and is therefore exempt from formal ethics review. The study collected no personal health or sensitive individual data and fell outside the National Commission on Informatics and Liberty declaration requirements (CNIL MR-004). Adherence to CNRS ethical best practices was respected by design as participation was voluntary and could be withdrawn at any time. Organisational affiliations are reported in aggregate only; no individual respondents are identified, and all data are stored securely with access restricted to the research team.

## 3. Results

A total of 35 actors responded to the survey from a total of 110 (32% response rate). The stakeholder distribution was healthcare professionals (n=18, 51.4% of respondents), patients and caregivers (n=6, 17.1%), academia (n=7, 20.0%), policymakers (n=3, 8.6%) and the pharmaceutical industry (n=1, 2.9%). Depending on the topic, response patterns varied across stakeholder categories. Most responses from healthcare professionals, academia and the single pharmaceutical industry respondent fell within “Strongly agree” and “Agree” categories. Patient representatives showed the greatest variability, with 26% of their responses in “Disagree” or “Strongly disagree” categories. Policymakers recorded the highest proportion of “Neutral” responses (37%). The sample includes representation across all intended stakeholder categories and captures perspectives from national, regional, and local levels of governance and practice.

### 3.1. Knowledge, Training, and Information Gaps

High consensus emerged across most stakeholder groups regarding substantial knowledge deficits about deprescribing. Among healthcare professionals, 83.3% reported familiarity with the deprescribing concept, but only 33.3% received training on it. This gap between awareness and competency-building was even more pronounced among policymakers. Only 33.3% were familiar with deprescribing, and only 33.3% had received training. Patient representatives showed the lowest knowledge levels, with 83.3% unfamiliar with the term “deprescribing” and 100% reporting no exposure to education or information campaigns on the topic.

Knowledge of specific deprescribing tools and guidelines was limited across groups. Among healthcare professionals, only the START/STOPP and EMPOWER tools were recognised by significant proportions of respondents. Among respondents, there was a general preference for French-validated resources, particularly HAS guidelines, CEPRIM and IATROPREV methodologies, and lists such as the Laroche criteria for potentially inappropriate medications. Academics were more likely to reference international protocols, including those from the Canadian ReCaD network, Australian Deprescribing Network and U.S. Deprescribing Network, alongside French resources.

Regarding information quality and availability, healthcare professionals expressed high consensus (94.4%) that clear, evidence-based protocols would facilitate deprescribing implementation. They also reported challenges accessing integrated and updated information systems. 72.2% agreed that better access to clinical decision support tools would help. Policymakers expressed concern about research data quality and robustness (66.7%), suggesting that stronger evidence could strengthen the policy case for deprescribing. Academics identified barriers to accessing real-world data for research (57.2%) and emphasised the need for modern digital platforms to support both research and clinical decision-making (57.2%).

Patients and caregivers demonstrated high consensus (100%) that having more information would positively impact their willingness to consider deprescribing. They specifically requested clear explanations of risks and benefits, simple instructions, and trustworthy sources, preferably from health authorities rather than pharmaceutical companies. 66.7% of patient representatives indicated that current information about medications is insufficient and unclear, and a similar proportion reported difficulty discussing health and medications with healthcare providers.

### 3.2. Attitudes, Values, and Trust

Stakeholder attitudes toward deprescribing are complex and divergent. Healthcare professionals showed high consensus (88.3%) in thinking that deprescribing is an important aspect of high-quality care, with 94.4% advocating for more strategies to be developed. This suggests substantial professional motivation to advance the practice and could serve as a foundation for policy initiatives. However, professional confidence in implementing deprescribing was more ambivalent. 83.3% reported some level of comfort discussing deprescribing with patients, but uncertainty emerged about proposing and managing deprescribing plans (66.7% agree, but no high consensus). Healthcare professionals were particularly concerned about potential negative patient outcomes (44.4% agree these concerns exist) and legal liability (66.7% agree this is a barrier), reflecting a moderate consensus on risk perceptions.

Among policymakers, there was moderate consensus (66.7%) supporting the link between deprescribing and its alignment with public health objectives. Similar agreement levels pointed to the incorporation of deprescribing into national policy frameworks. However, there was no clear consensus among policymakers on whether France’s current regulatory framework is prepared to support the implementation of deprescribing protocols.

The single pharmaceutical industry respondent acknowledged that deprescribing aligns with public health goals and sustainable healthcare and agreed that sufficient evidence exists to support its implementation. While not representative, it is noteworthy that this respondent also indicated that deprescribing could represent an economic opportunity to focus on innovative therapies rather than traditional medication volumes.

Patient attitudes revealed important nuances. There was high consensus (100%) on their perceived value for medications, with 83.3% stating that they feel their health is more valued when offered an alternative medication rather than reducing or discontinuing it. However, patients also showed high consensus (83.4%) when faced with questions about trusting their healthcare team regarding medication decisions, and 66.7% understood the importance of reducing the number of medications they were taking.

Regarding priority populations, results showed a moderate consensus (66.7%) among healthcare professionals that deprescribing is more important for elderly patients than younger adults. However, academics and some professionals noted that deprescribing principles apply across the life course and that considering age as a defining factor might limit deprescribing scope and efforts.

### 3.3. Resistance, Communication, and Collaboration

There was high consensus (83.3%) among healthcare professionals regarding the patient resistance they experienced when proposing deprescribing plans. This perception aligns with patient resistance to medication changes (66.7%), as they feel their health would deteriorate without medications or believe that deprescribing practices are a form of under-provision of health. Despite this, there is a high consensus among patients that they trust healthcare professionals (83.4%) and value having discussions around their medications.

Communication barriers exist at multiple levels. There was high consensus among healthcare professionals (72.2%) regarding a lack of time to engage in conversations with patients, to participate in medication reviews or in collaborative decision-making processes. Professionals reported that this affected resistance since patient engagement is necessary for successful communication and understanding. In fact, our results showed that patients have difficulties discussing their health status and medications with healthcare professionals (66.7%). Healthcare professionals emphasised that effective communication requires time for explaining risks and benefits, exploring patient values and preferences, ensuring long-term patient monitoring and building coordination plans among specialists.

Interprofessional and intersectoral collaboration emerged as key challenges. Healthcare professionals reported high consensus (83.4%) that collaboration is fundamental for achieving better deprescribing outcomes, recognising that working collaboratively in France is difficult. They identified specific collaboration needs, including better integration between primary care settings in the community and hospital, the incorporation of advanced practice roles at the national level, the creation of multidisciplinary team models, and the development of interoperable communication tools that enable the efficient sharing of medical records and prescription data.

Policymakers and healthcare professionals showed strong agreement (66.6% and 88.9%, respectively) that intersectoral collaboration is essential to incentivise deprescribing and develop effective national strategies. Academics outlined, with moderate consensus (57.2%), that the lack of interdisciplinary collaboration limits scope and impact in the field. Academics noted additional barriers specific to research and knowledge production, including publication bias favouring drug trials over deprescribing studies (71.5%), limited interdisciplinary research networks (57.2%), and challenges disseminating findings effectively to policy and practice audiences (85.8%). They also highlighted that these barriers affect the evidence base available to guide clinical and policy decisions.

### 3.4. Resources: Tools, Data, and Time

While resource constraints emerged as a domain having general cross-stakeholder consensus, there was divergence when it came to which resources were considered most critical and for whom. Healthcare professionals expressed high consensus (72.2%) that time constraints were a major limitation preventing more frequent exploration of deprescribing options. They agreed that limited consultation time affects their ability to review complex medication regimes, engage patients in shared decision-making, coordinate with other specialists, and monitor outcomes after medication changes. Interestingly, policymakers did not identify time constraints as a significant barrier.

Access to quality tools, protocols, and data showed significant cross-stakeholder consensus. Healthcare professionals agreed that clear, evidence-based protocols (94.4%) and clinical decision support tools (72.2%) would facilitate a wider implementation of deprescribing. Policymakers emphasised the need for robust, data-driven research to strengthen the policy case (66.7%), and academics highlighted challenges accessing real-world data for research (57.2%) and the value of digital platforms for both research and clinical support (57.2%). However, perspectives diverged on what constitutes adequate tools. Healthcare professionals prefer French-validated protocols (HAS guidelines, CEPRIM, IATROPREV, Laroche list) and emphasise the value of clinical experience and in-house knowledge developed by multidisciplinary teams. Patients emphasised the need for accessible, plain-language materials explaining medication risks and benefits.

Concerns about data quality and liability emerged among professionals and policymakers. Healthcare professionals worried about negative patient outcomes (44.4%) and legal repercussions (66.7%) associated with deprescribing. They believe that most current protocols and data are not adequate to protect them. Additionally, policymakers highlighted concerns around research robustness (66.7%), suggesting that expanding the evidence base could reduce reluctance.

Human resource constraints also emerged in our open-ended questions, with responses including insufficient pharmacist integration in primary care medication reviews, limited nursing staff available for patient monitoring and education, and the absence of psychologists and social workers to support patients through medication changes that may affect mental health or require behavioural adaptations.

### 3.5. Policy, Regulation, and Institutional Support

This domain revealed the greatest divergence between stakeholder groups. For healthcare professionals, there was high consensus (88.9%) that current reimbursement policies and payment methods provide insufficient incentives to promote deprescribing. While linking deprescribing to financial incentives was considered inappropriate, the use of quality metrics to encourage proper medication use was viewed as more appropriate.

Policymakers were more positive about incentives, with moderate consensus (66.6%) for financial incentives and creative reimbursement policies to promote deprescribing. However, they also acknowledged bureaucratic delays in policy implementation (66.7%) and concerns around the pharmaceutical lobby (66.7%) as impediments to advancing the topic. Regarding regulatory preparedness, there was no clear consensus among policymakers on whether France’s regulatory framework could support deprescribing protocols.

Academics were more critical, with moderate to high consensus (71.5%) that political will to advance deprescribing is insufficient, with similar agreement (71.4%) regarding the French policy environment as reluctant to embrace innovative approaches quickly. They also identified insufficient institutional support for deprescribing research (71.5%) and inadequate funding (71.5%) as barriers for evidence generation. Academics were uncertain about stimuli for research on deprescribing, and showed scepticism that funding alone, including more targeted grants, could overcome systemic obstacles.

On the pharmaceutical industry side, our respondent indicated that new regulatory frameworks for deprescribing would affect product portfolios and require business strategy adjustments. Despite this, the opinion is that industry is well-positioned to adapt and innovate, particularly by focusing on novel therapies rather than traditional medication volumes. The respondent also highlighted that deprescribing could open new economic opportunities.

Regarding institutional support structures, healthcare professionals and patients identified several important actors, including CPAM, ARS, OMEDIT, private insurance companies (“*mutuelles*”), and risk management organisations. Professionals believed *mutuelles* are well-positioned to incentivise to act on quality and to prevent over-prescribing. Patients emphasised the need for concrete services from the institutions, such as counselling, public health campaigns, more multidisciplinary support teams, and accessible educational materials.

### 3.6. Opportunities: Clinical, Economic, and Environmental Benefits

Perceived opportunities associated with deprescribing showed relatively strong cross-stakeholder consensus, particularly regarding clinical benefits and environmental sustainability.

Clinical and quality of life benefits garnered the highest agreement. Healthcare professionals reported high consensus that deprescribing improves patient safety (100%) and quality of life (88.8%). Academia showed a similar strong agreement (85.7%) on these topics. Policymakers concurred regarding improvements in safety (66.6%) and quality of life (66.6%), though with moderate rather than high consensus, with even the pharmaceutical industry representative acknowledging these benefits.

Economic impacts showed more complex patterns. Healthcare professionals (94.5%) and policymakers (66.7%) agreed that deprescribing contributes to healthcare system cost savings, showing moderate consensus (72.2% and 66.7%, respectively) around the long-term economic benefits justifying any short-term implementation costs. Academia showed moderate consensus (57.1%) that research demonstrates clear cost-effectiveness, though academics were divided on whether deprescribing reduces overall healthcare expenditure. Patients believed that stopping unnecessary medications would reduce their personal healthcare expenses (66.7%), but an equal proportion highlighted that economic considerations alone would not be enough to increase their willingness to reduce medications. The pharmaceutical industry highlighted the new economic opportunities around the implementation of deprescribing mechanisms, though they also acknowledged market share concerns and financial stress in the absence of incentives in this sector.

Environmental sustainability emerged as a surprising area of relatively strong consensus. Healthcare professionals expressed high consensus that deprescribing reduces pharmaceutical waste (88.9%) and the environmental impact of medicines (88.9%). Policymakers generally agreed on the reduction of pharmaceutical waste (66.7%) by optimising the use of medicines (66.7%). Academia showed moderate consensus (57.2%) on deprescribing and the positive environmental impacts, but a strong agreement (85.7%) in the link between deprescribing research and waste reduction strategies. Even patients agreed (66.7%) that reducing unnecessary medications would benefit the environment, though they were less certain about contributions to healthcare system sustainability. Cross-stakeholder consensus also emerged regarding the importance of developing robust policies linking deprescribing and environmental sustainability. Professionals (94.4%), academics (100%), and policymakers (66.7%) all agreed that this is important for France. This is particularly notable given that environmental considerations in healthcare policy are relatively recent in France. The pharmaceutical sector also agreed that deprescribing aligns with sustainable healthcare but showed uncertainty about specific environmental metrics.

## 4. Discussion

The findings from this comprehensive mapping of stakeholder perspectives on deprescribing across France’s health policy ecosystem align with international literature on barriers and enablers and highlight patterns shaped by the institutional and cultural context in France. Patterns around knowledge gaps among professionals and patients (Farrell et al., 2023; Reeve, 2020; van Poelgeest et al., 2022), communication issues and patient resistance (Assurance Maladie, 2025; Barbara Starfield et al., 2005; Groupe Onepoint, 2021; Océane Degang, 2022; Peat et al., 2022; Rodwin, 2020; Tomaschek et al., 2022; Turner et al., 2018, 2020) and resource constraints (Barbara Starfield et al., 2005; Groupe Onepoint, 2021) are widely acknowledged in the literature. However, in the French context, some interesting particularities include a preference for using national protocols over international guidance and the challenges associated with improving interprofessional collaborations. These may reflect historic patterns that favour centralised approaches, and an institutional configuration that highly protects physician autonomy and independent practice models (Océane Degang, 2022; Rodwin, 2020; Tomaschek et al., 2022). Furthermore, the low response rate from the policy ecosystem might be a reflection of France’s technocratic governance, where officials are usually cautious about expressing positions in public that could constitute a future commitment (Simonet, 2021).

A recent comparative analysis that included stakeholder consultations in Ireland and Canada revealed that, similar to France, Irish stakeholders emphasised the need for multidisciplinary collaboration and clearer guidelines. However, because its system is less fragmented, governance appeared to enable more aligned views between policymakers and professionals. Canadians reported stronger patient engagement and advocacy for deprescribing, which can be a reflection of increased patient involvement in shaping health policies in Canada compared to France (Riordan et al., 2016; Tannenbaum et al., 2014; Wilkinson et al., 2017).

The use of an adapted Policy Delphi approach revealed distinct patterns of consensus and divergence that can contribute to identifying stakeholder alignment. This enables the identification of topics conducive to negotiation dialogues that can inform policy-making interventions (Landeta, 2006; Pauline Van Ngoc, 2025). This aligns with principles of collaborative governance, which emphasise the importance of understanding stakeholder positions as a precursor to effective multi-actor coordination (Ansell & Gash, 2008). In a context of de-implementation strategies, this approach can also be combined with other tools, such as the recent “Policy Lever Inventory for Low-Value Care” (Warkentin et al., 2026) to inform evidence-based policy.

The three domains that emerged as having high cross-stakeholder consensus can be thought of as immediate policy opportunities. The first is regarding knowledge and training gaps, which point to the need to implement educational interventions. Moreover, consensus around the clinical benefits of deprescribing for patient safety and quality of life also invites specific actions from civil society, patient advocacy groups and healthcare professional associations. Finally, agreement on the impact of deprescribing on environmental sustainability offers a novel policy framing that aligns with the green agenda and the promotion of eco-responsible interventions (Barratt et al., 2022).

On the other hand, while professionals and academics see major gaps in policy and regulatory readiness, the unclear position of policymakers requires more extended negotiations. For example, the role of incentives, especially economic ones, remains unclear. Most professionals resist purely economic motivations, whereas policymakers and the pharmaceutical industry remain open to discussion and exploration of alternatives.

### Implications for practice: the policy Delphi heatmap as a governance tool

This study provides a methodological innovation by adapting the Policy Delphi framework as a tool for stakeholder and governance analysis in healthcare reform. Traditional approaches to stakeholder engagement in health policy often use generic consultation processes for opinions but fail to systematically assess consensus patterns or identify strategic intervention points (Abelson et al., 2007; Mitton et al., 2009). This continues to occur despite results highlighting the importance of structured approaches to knowledge mobilisation that map stakeholder positions to facilitate negotiation and reach success (Cairney & Oliver, 2017; Lavis et al., 2009). Our heatmap provides a visual representation of policy traction and friction. This is particularly valuable in a fragmented governance context, such as in France, where coordination across multiple institutional actors is essential but challenging (Dominique Polton et al., 2021; Simonet, 2023).

This approach enables four governance functions. First, it eases strategic prioritisation for policymakers by identifying high-consensus domains and identifies interventions that can build momentum through early wins before diving into more complex issues. As a result of this study, for example, the focus could be on implementing training initiatives and environmental sustainability interventions. Second, this approach helps to build coalitions by identifying which stakeholders share similar perspectives so they can develop strategic alliances. In our case, coalitions could be formed to advocate for improved communication and information. Third, this approach supports negotiations, for example, around policy and regulation, as it helps policymakers to understand divergent issues that require dialogue and compromise to reach a solution instead of relying on top-down decrees. Finally, this approach provides a learning framework: by performing periodic re-assessments, it can help track how stakeholder perspectives evolve over time as new policies are implemented, creating feedback loops for adaptive governance (SciencesPo & LIEPP, 2024).

### Limitations and Future Research

This study has several limitations that contextualise findings and indicate directions for further research. The 32% response rate and low participation from policymakers and the pharmaceutical industry limit generalisability within those stakeholder categories. The perspectives of the only respondent from the pharmaceutical industry, while included for completeness, cannot represent the diversity of opinions within the pharmaceutical sector. These groups are difficult to engage in academic research due to time constraints, institutional protocols, and potential sensitivity around policy positions (Jill Pearcy, 2024; Oliver et al., 2014; Tricco et al., 2018). Future research employing targeted interviews with policymakers and industry representatives would enrich our understanding of what they consider important for decision-making.

Patient representatives were organisations rather than individual patients or caregivers. While patient organisations and advocacy groups play important roles in French health policy, their perspectives may not fully capture the experiences and views of typical patients managing medications. Participatory research methods, such as patient advisory panels or co-design could complement these perspectives (Bate & Robert, 2006).

The one-round survey design, while appropriate for exploratory mapping, does not enable the iterative refinement and consensus-building of multi-round Delphi studies. The ongoing DEPREFLEX program, of which this exploratory study was a part of, can address this issue in future research (Etienne Nouguez et al., n.d.). Finally, this study captures perspectives at a single time point, but stakeholder views evolve as the policy debate progresses and pilot programs generate evidence, both at the national and international scale. Longitudinal tracking of stakeholder perspectives would help to understand these dynamics and support adaptive policy development.

Despite these limitations, this study provides essential baseline mapping of the French deprescribing policy landscape and demonstrates the value of Policy Delphi approaches for governance analysis in complex healthcare reform.

## 5. Conclusions and Recommendations

Deprescribing represents a critical yet underdeveloped health policy priority in France. This study reveals that there exists consensus on core values, including patient safety, quality of life, and environmental sustainability that could be the germ for a broader national deprescribing framework. However, divergent perspectives on policy instruments, regulatory readiness, and institutional support indicate that advancing deprescribing requires more than technical intervention. This study points to building strategic coalitions, structured stakeholder dialogue, governance coordination and iterative policy learning as fundamental tools to advance on this topic in the future. Furthermore, the Policy Delphi heatmap provides a novel governance tool for the identification of stakeholder alignment, immediate action points, or a further need for negotiation, and has proven to be a useful tool for complex health reforms requiring multi-actor coordination.

We suggest actions based on consensus levels first, from immediate actions on high-consensus domains, such as training and environmental framing, to structured dialogue and pilotage for moderate-consensus areas, including collaboration models and communication tools. Policy tools such as incentives and regulation could be implemented following sustained negotiation on low-consensus areas. These differentiated approaches build on the idea that policy progress does not need complete consensus but rather a strategic approach to build momentum. We have conceptualised such a national deprescribing framework in Figure 1.

**Figure 1:**
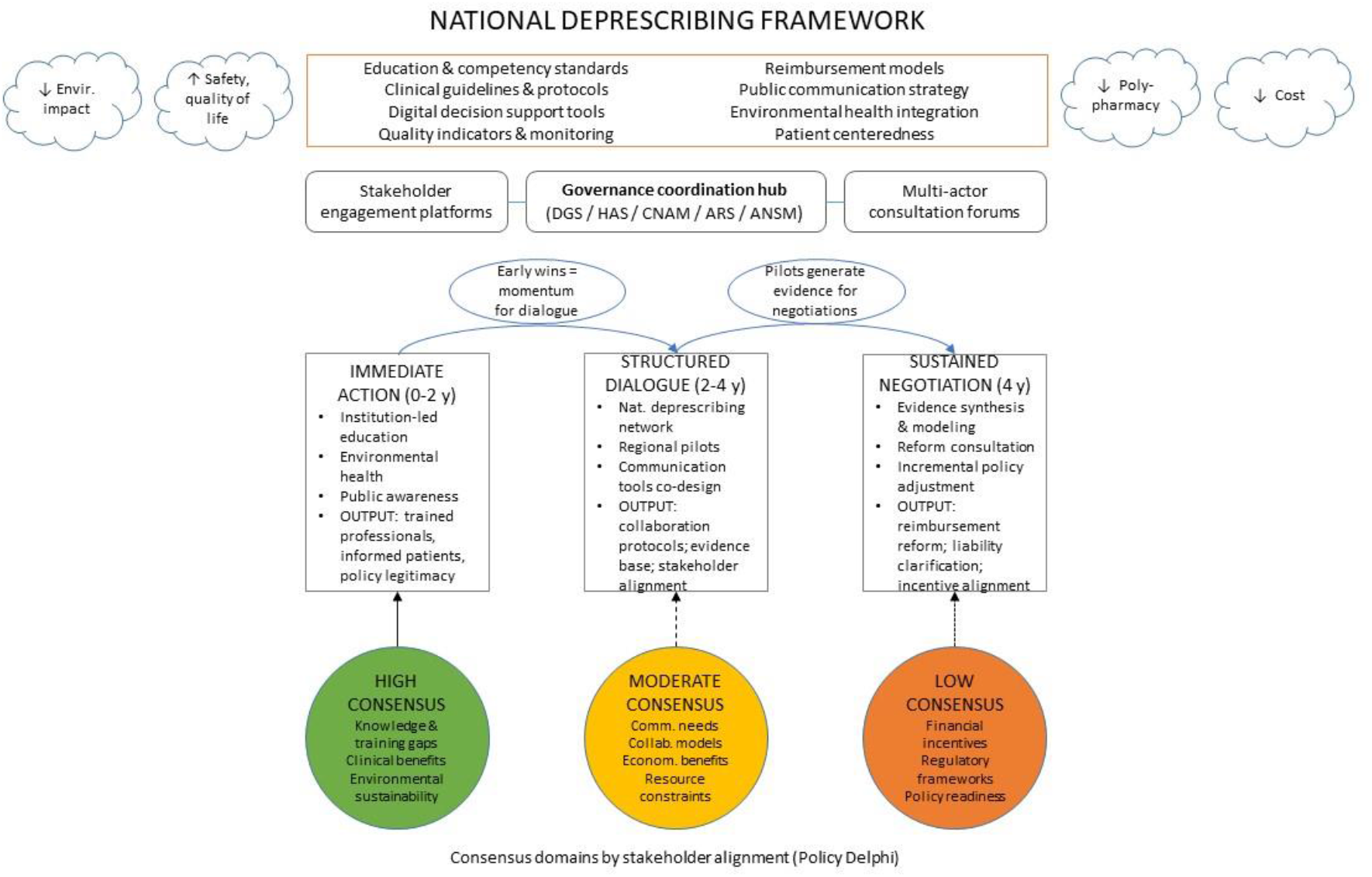
From Consensus to Coordination - Conceptual Model for A National Deprescribing Framework. *Note: Policy progress does not require complete stakeholder consensus. Strategic sequencing capitalizes on areas of alignment to build momentum, while creating structured spaces for negotiation on contested issues. The pathway from consensus mapping to coordinated action requires intentional governance—convening diverse actors, generating shared evidence, and translating alignment into institutional change*.

FIGURE 1 HERECross-stakeholder recognition of the benefits of deprescribing is a start, but translating this into systematic implementation requires moving beyond fragmented initiatives to more coordinated policy frameworks. As France develops its next health policy agenda, deprescribing is an opportunity to demonstrate that clinical quality, fiscal responsibility, and environmental protection are not competing priorities and can work synergistically. This requires the political will to convene diverse stakeholders, the institutional capacity to coordinate across fragmented governance structures, and the strategy to navigate around consensus and divergence (see Box 1).

### Box 1

Strategic recommendations

- **Launch a National Deprescribing Education Initiative:** Develop and validate deprescribing competency frameworks for different professional groups and coordinate their integration into health professional education curricula. This initiative should include public-facing components, including patient education materials and mass media campaigns leveraging the capacities of patient organisations and advocacy groups.
- **Foster and promote the work of the French National Deprescribing Network** (currently in progress): Co-design implementation frameworks, develop communication and shared decision-making protocols, collaborative medication review models, a competency framework for new healthcare professional roles, and monitoring indicators. In collaboration with parallel work on “green” prescribing (Bounoure et al., 2025), integrate deprescribing into a broad sustainability agenda by co-creating a roadmap between key stakeholders that focuses on the development of eco-prescribing guidelines and responsible medication.
- **Commission comprehensive evidence synthesis on policy instruments and incentives:** Incentivise research priorities including economic modelling of alternative reimbursement structures for medication review services, comparative policy analysis examining deprescribing frameworks in similar healthcare systems, a legal analysis of liability issues and potential regulatory reforms to enable expanded professional roles, and implementation science studies documenting effective incentives and supports to inform the National Deprescribing Network.

## Data Availability

All data produced in the present study are available upon reasonable request to the authors.

## Acknowledgments

The authors wish to thank all participants of a one-day workshop on deprescribing policies held at Sciences Po, Paris, on Dec 4, 2024, for their valuable insight and sustained support. This article has received support from the French National Research Agency (ANR) *Investissement d’avenir* program as part of the IdEx Université Paris Cité (ANR-18-IDEX-0001).

---

**Table 1:** Stakeholder Characteristics and Response Rates.

| Stakeholder Group | Organizations Invited (n) | Responses Received (n) | Response Rate (% of invited) | Distribution of Respondents (% of all) |
| --- | --- | --- | --- | --- |
| Healthcare Professionals | 45 | 18 | 40.0% | 51.4% |
| Patients & Caregivers | 22 | 6 | 27.3% | 17.1% |
| Academia & Research | 18 | 7 | 38.9% | 20.0% |
| Policymakers & Regulators | 20 | 3 | 15.0% | 8.6% |
| Pharmaceutical Industry | 5 | 1 | 20.0% | 2.9% |
| **Total** | **110** | **35** | **31.8%** | **100%** |

**Note:** Response rates calculated as percentage of invited organizations within each stakeholder category that provided completed surveys. Distribution of respondents shows each group's proportion of the total sample (n=35).

**Table 2:** Summary of Consensus Strength Across Policy Domains.

| Policy Domain | Healthcare Professionals | Patients & Caregivers | Academia & Research | Policymakers & Regulators | Pharmaceutical Industry | Cross-Stakeholder Pattern |
| --- | --- | --- | --- | --- | --- | --- |
| Knowledge & Training | High | High | High | Moderate | N/A | Strong Convergence |
| Attitudes & Values | High | Moderate | High | Moderate | N/A | Moderate Convergence |
| Patient Resistance | High | High | Low | No consensus | N/A | Partial Convergence |
| Communication & Collaboration | High | Moderate | Moderate | Moderate | N/A | Moderate Convergence |
| Resources (time, tools, data) | High | Moderate | Moderate | Low | N/A | Strong Divergence |
| Policy & Regulatory Support | High | Moderate | Moderate | No consensus | N/A | Strong Divergence |
| Clinical Benefits | High | High | High | Moderate | N/A | Strong Convergence |
| Economic Opportunities | High | Low | Moderate | Moderate | N/A | Moderate Convergence |
| Environmental Benefits | High | Moderate | Moderate | Moderate | N/A | Strong Convergence |

**Legend: Cells represent convergence within stakeholder groups except for the “cross stakeholder pattern”**.

- **High:** ≥70% agreement in single category OR ≥80% in grouped categories
- **Moderate:** ≥60% and <70% in single category OR ≥70% and <80% in grouped categories
- **Low:** >50% and <60% in single category OR ≥60% and <70% in grouped categories
- **No consensus:** Agreement below thresholds
- **(-):** Consensus indicates *lack* or *insufficiency* (e.g., insufficient knowledge, inadequate support)
- **(+):** Consensus indicates endorsement or recognition of benefits
- **No data:** Insufficient responses to assess consensus

**Cross-Stakeholder Patterns:**

- **Strong Convergence:** High/moderate consensus across ≥4 stakeholder groups with similar direction
- **Moderate Convergence:** Moderate consensus across ≥3 groups with similar direction
- **Partial Convergence:** Consensus in 2-3 groups, divergent or absent in others
- **Strong Divergence:** Opposing consensus directions or mix of high consensus and no consensus across groups

## Footnotes

1 Term used to encapsulate the broad overuse or misuse of care that negatively impacts both patients and healthcare systems.

## References

Ministère des Solidarités et de la Santé. (2023). Projet Stratégie nationale de santé 2023-2033 (Projet soumis à consultation).

Abelson, J., Giacomini, M., Lehoux, P., & Gauvin, F.-P. (2007). Bringing ‘the public’ into health technology assessment and coverage policy decisions: From principles to practice. Health Policy, 82(1), 37–50. 10.1016/j.healthpol.2006.07.009

Ailabouni, N. J., Reeve, E., Helfrich, C. D., Hilmer, S. N., & Wagenaar, B. H. (2022). Leveraging implementation science to increase the translation of deprescribing evidence into practice. Research in Social and Administrative Pharmacy, 18(3), 2550–2555. 10.1016/j.sapharm.2021.05.018

Ansell, C., & Gash, A. (2008). Collaborative Governance in Theory and Practice. Journal of Public Administration Research and Theory, 18(4), 543–571. 10.1093/jopart/mum032

Assurance Maladie. (2025). Mon espace santé a 3 ans et déjà plus de 2,5 millions d’utilisateurs tous les mois. In https://sante.gouv.fr/actualites/presse/communiques-de-presse/.

Barbara Farrell, Kevin Pottie, Wade Thompson, Taline Boghossian, Lisa Pizzola, Farah Joy Rashid, Carlos Rojas-Fernandez, Kate Walsh, Vivian Welch, & Paul Moayyedi. (2017). Deprescribing proton pump inhibitors. Evidence-based clinical practice guideline. Canadian Family Physician - Le Médecin de Famille Canadien, 61.

Barbara Starfield, Leiyu Shi, & James Macinko. (2005). Contribution of Primary Care to Health Systems and Health. The Milbank Quarterly, 83(3), 457–502. 10.1111/j.1468-0009.2005.00409.x

Barratt, A. L., Bell, K. J., Charlesworth, K., & McGain, F. (2022). High value health care is low carbon health care. Medical Journal of Australia, 216(2), 67–68. 10.5694/mja2.51331

Bate, P., & Robert, G. (2006). Experience-based design: from redesigning the system around the patient to co-designing services with the patient. Quality and Safety in Health Care, 15(5), 307–310. 10.1136/qshc.2005.016527

Bolt, J., Abdoulrezzak, R., & Inglis, C. (2023). Barriers and enablers to deprescribing of older adults and their caregivers: a systematic review and meta-synthesis. European Geriatric Medicine, 14(6), 1211–1222. 10.1007/s41999-023-00879-7

Bounoure, F., Dupray, S., Wils, J., Taillemite, S., Brunn, M., Siefridt, C., Le Clech, N., Réveillon-Istin, M., Skiba, M., Bouglé, C., & Lahiani-Skiba, M. (2025). L’éco-prescription, une nouvelle compétence majeure pour les professionnels de santé. Annales Pharmaceutiques Françaises. 10.1016/j.pharma.2025.10.003

Bras, P.-L. (2008). Organisation des soins et régulation des dépenses ambulatoires : le rôle des syndicats médicaux. Les Tribunes de La Santé, n° 18(1), 47–56. 10.3917/seve.018.0047

Brunn, M., & Hassenteufel, P. (2021). France. In Health Politics in Europe (pp. 558–589). Oxford University PressOxford. 10.1093/oso/9780198860525.003.0025

Cairney, P., & Oliver, K. (2017). Evidence-based policymaking is not like evidence-based medicine, so how far should you go to bridge the divide between evidence and policy? Health Research Policy and Systems, 15(1), 35. 10.1186/s12961-017-0192-x

Carollo, M., Boccardi, V., Crisafulli, S., Conti, V., Gnerre, P., Miozzo, S., Omodeo Salè, E., Pieraccini, F., Zamboni, M., Marengoni, A., Onder, G., Trifirò, G., Antonioni, R., Selleri, M., Vitturi, G., Filippelli, A., Corrao, S., Medea, G., Nobili, A., … Veronese, N. (2024). Medication review and deprescribing in different healthcare settings: a position statement from an Italian scientific consortium. Aging Clinical and Experimental Research, 36(1), 63. 10.1007/s40520-023-02679-2

Charbonneau, M., Morgan, S. G., Gagnon, C., Sadowski, C. A., Silvius, J. L., Tannenbaum, C., & Turner, J. P. (2024). Factors influencing the effects of policies and interventions to promote the appropriate use of medicines in high-income countries: A rapid realist review. Health Policy, 142, 105027. 10.1016/j.healthpol.2024.105027

Cross, A. J., Etherton-Beer, C. D., Clifford, R. M., Potter, K., & Page, A. T. (2021). Exploring stakeholder roles in medication management for people living with dementia. Research in Social and Administrative Pharmacy, 17(4), 707–714. 10.1016/j.sapharm.2020.06.006

Daunt, R., Curtin, D., & O’Mahony, D. (2023). Polypharmacy stewardship: a novel approach to tackle a major public health crisis. The Lancet Healthy Longevity, 4(5), e228–e235. 10.1016/S2666-7568(23)00036-3

Doherty, A. J., Boland, P., Reed, J., Clegg, A. J., Stephani, A.-M., Williams, N. H., Shaw, B., Hedgecoe, L., Hill, R., & Walker, L. (2020). Barriers and facilitators to deprescribing in primary care: a systematic review. BJGP Open, 4(3), bjgpopen20X101096. 10.3399/bjgpopen20X101096

Dominique Polton, Hélène Chaput, Mickaël Portela, Quentin Laffeter, & Christelle Millien. (2021). Remédier aux pénuries de médecins dans certaines zones géographiques. Les leçons de la littérature internationale.

DRESS. (n.d.). Les dépenses de santé en 2023. Resultats de comptes de la santé. Édition 2024.

Drusch, S., Zureik, M., & Herr, M. (2023). Potentially inappropriate medications and polypharmacy in the older population: A nationwide cross-sectional study in France in 2019. Therapies, 78(5), 575–584. 10.1016/j.therap.2023.02.001

Etienne Nouguez, Jonathan Siscic, & Matthias Brunn. (n.d.). Déprescription et perceptions - Une approche mixte pour comprendre les freins et leviers (DEPREFLEX). Https://Www.Sciencespo.Fr/Liepp/Fr/Recherche/Projet/Deprescription-et-Perceptions-Une-Approche-Mixte-Pour-Comprendre-Les-Freins-et-Leviers-Depreflex/.

Farrell, B., Raman-Wilms, L., Sadowski, C. A., Mallery, L., Turner, J., Gagnon, C., Cole, M., Grill, A., Isenor, J. E., Mangin, D., McCarthy, L. M., Schuster, B., Sirois, C., Sun, W., & Upshur, R. (2023). A Proposed Curricular Framework for an Interprofessional Approach to Deprescribing. Medical Science Educator, 33(2), 551–567. 10.1007/s40670-022-01704-9

Forest, E., Ireland, M., Yakandawala, U., Cavett, T., Raman-Wilms, L., Falk, J., McMillan, D., Linthorst, R., Kosowan, L., Labine, L., & Leong, C. (2021). Patient values and preferences on polypharmacy and deprescribing: a scoping review. International Journal of Clinical Pharmacy, 43(6), 1461–1499. 10.1007/s11096-021-01328-w

François Buton. (2024). Une élite à part. La politique comme déplacement des médecins dans le champ du pouvoir. In F. Buton (Ed.), En déplacement. Le passage des frontières professionnelles en question. (pp. 207–223). ENS Éditions. 10.4000/12h9n

Fraser, G. R. L., Lambooij, M. S., van Exel, J., Ostelo, R. W. J. G., van Harreveld, F., & de Wit, G. A. (2024). Factors associated with patients’ demand for low-value care: a scoping review. BMC Health Services Research, 24(1), 1656. 10.1186/s12913-024-12093-7

Gillespie, R. J., Harrison, L., & Mullan, J. (2018). Deprescribing medications for older adults in the primary care context: A mixed studies review. Health Science Reports, 1(7). 10.1002/hsr2.45

Gouvernement de France. (2023). France Nation Verte: Planification écologique du système de santé.

Groupe Onepoint. (2021). La santé des Français et l’hôpital au cœur de la campagne présidentielle. In https://www.groupeonepoint.com/fr/actualites/.

Heinrich, C. H., Hurley, E., McCarthy, S., McHugh, S., & Donovan, M. D. (2022). Barriers and enablers to deprescribing in long-term care facilities: a ‘best-fit’ framework synthesis of the qualitative evidence. Age and Ageing, 51(1). 10.1093/ageing/afab250

INSEE. (2025). Tableau de bord de l’économie française – Population.

Jill Pearcy. (2024). Why it matters to industry reputation and what we can we do about it. In https://www.abpi.org.uk/media/blogs/2024/march/.

Kanger, L., Sovacool, B. K., & Noorkõiv, M. (2020). Six policy intervention points for sustainability transitions: A conceptual framework and a systematic literature review. Research Policy, 49(7), 104072. 10.1016/j.respol.2020.104072

Khazzaka, M. (2019). Pharmaceutical marketing strategies’ influence on physicians’ prescribing pattern in Lebanon: ethics, gifts, and samples. BMC Health Services Research, 19(1), 80. 10.1186/s12913-019-3887-6

Landeta, J. (2006). Current validity of the Delphi method in social sciences. Technological Forecasting and Social Change, 73(5), 467–482. 10.1016/j.techfore.2005.09.002

Laroche, M.-L., Roux, B., & Grau, M. (2017). Iatrogénie médicamenteuse chez la personne âgée, comprendre et agir. Actualités Pharmaceutiques, 56(571), 28–32. 10.1016/j.actpha.2017.09.023

Lavis, J. N., Oxman, A. D., Lewin, S., & Fretheim, A. (2009). SUPPORT Tools for evidence-informed health Policymaking (STP). Health Research Policy and Systems, 7(S1), I1. 10.1186/1478-4505-7-S1-I1

Matthias Brunn, Karen Berg Brigham, Karine Chevreul, & Cristina Hernández-Quevedo. (2018). The impact of the crisis on the health system and health in France. In Economic crisis, health systems and health in Europe: Country experience (pp. 283–321).

Mitton, C., Smith, N., Peacock, S., Evoy, B., & Abelson, J. (2009). Public participation in health care priority setting: A scoping review. Health Policy, 91(3), 219–228. 10.1016/j.healthpol.2009.01.005

Océane Degang. (2022). La collaboration entre les Médecins Généralistes et les Sages-Femmes libérales au sein des maisons de santé pluriprofessionnelles.

OECD. (2023). France: Country Health Profile 2023. 10.1787/07c48f9f-en

Okeowo, D. A., Zaidi, S. T. R., Fylan, B., & Alldred, D. P. (2023). Barriers and facilitators of implementing proactive deprescribing within primary care: a systematic review. International Journal of Pharmacy Practice, 31(2), 126–152. 10.1093/ijpp/riad001

Oliver, K., Innvar, S., Lorenc, T., Woodman, J., & Thomas, J. (2014). A systematic review of barriers to and facilitators of the use of evidence by policymakers. BMC Health Services Research, 14(1), 2. 10.1186/1472-6963-14-2

OMEDIT Pays de la Loire. (n.d.). Quelles sont les principales missions de l’OMEDIT?

Pauline Van Ngoc. (2025). Understanding and improving benzodiazepines and Z-drugs management in primary care.

Peat, G., Fylan, B., Marques, I., Raynor, D. K., Breen, L., Olaniyan, J., & Alldred, D. P. (2022). Barriers and facilitators of successful deprescribing as described by older patients living with frailty, their informal carers and clinicians: a qualitative interview study. BMJ Open, 12(3), e054279. 10.1136/bmjopen-2021-054279

Perera, I. M. (2022). Interest group governance and policy agendas. Governance, 35(3), 869– 886. 10.1111/gove.12615

Reeve, E. (2020). Deprescribing tools: a review of the types of tools available to aid deprescribing in clinical practice. Journal of Pharmacy Practice and Research, 50(1), 98– 107. 10.1002/jppr.1626

Reeve, E., Gnjidic, D., Long, J., & Hilmer, S. (2015). A systematic review of the emerging definition of ‘deprescribing’ with network analysis: implications for future research and clinical practice. British Journal of Clinical Pharmacology, 80(6), 1254–1268. 10.1111/bcp.12732

Reeve, E., Thompson, W., & Farrell, B. (2017). Deprescribing: A narrative review of the evidence and practical recommendations for recognizing opportunities and taking action. European Journal of Internal Medicine, 38, 3–11. 10.1016/j.ejim.2016.12.021

Richard, A., Mariotti, B., Piñol-Domenech, N., Vorilhon, P., & Vaillant-Roussel, H. (2023). Prescription-free consultation in France and Europe: Rates’ evolution physicians’ and patients’ perceptions from 2005 to 2019, a systematic review. Therapies, 78(6), 733–741. 10.1016/j.therap.2023.02.011

Riordan, D. O., Walsh, K. A., Galvin, R., Sinnott, C., Kearney, P. M., & Byrne, S. (2016). The effect of pharmacist-led interventions in optimising prescribing in older adults in primary care: A systematic review. SAGE Open Medicine, 4. 10.1177/2050312116652568

Rodwin, M. A. (2020). Pharmaceutical Price and Spending Controls in France: Lessons for the United States. International Journal of Health Services, 50(2), 156–165. 10.1177/0020731419897580

SciencesPo, & LIEPP. (2024). Déprescrire dans une perspective santé-environnement: quelles pistes pour les politiques publiques en France?. In https://www.sciencespo.fr/liepp/fr/evenements/.

Silva Almodóvar, A., Keller, M. S., Lee, J., Mehta, H. B., Manja, V., Nguyen, T. P. P., Pavon, J. M., Terman, S. W., Hoyle, D., Mixon, A. S., & Linsky, A. M. (2024). Deprescribing medications among patients with multiple prescribers: A socioecological model. Journal of the American Geriatrics Society, 72(3), 660–669. 10.1111/jgs.18667

Simonet, D. (2021). French Idiosyncratic Health-Care Reforms, Performance Management and Its Political Repercussions. Risk Management and Healthcare Policy, Volume 14, 2971– 2981. 10.2147/RMHP.S306381

Simonet, D. (2023). Health Care Reforms, Power Concentration, and Receding Citizen Participation. Risk Management and Healthcare Policy, Volume 16, 1359–1364. 10.2147/RMHP.S421397

Tannenbaum, C., Martin, P., Tamblyn, R., Benedetti, A., & Ahmed, S. (2014). Reduction of Inappropriate Benzodiazepine Prescriptions Among Older Adults Through Direct Patient Education. JAMA Internal Medicine, 174(6), 890. 10.1001/jamainternmed.2014.949

The Shift Project. (2023). Décarboner la santé pour soigner durablement.

Tomaschek, R., Lampart, P., Scheel-Sailer, A., Gemperli, A., Merlo, C., & Essig, S. (2022). Improvement Strategies for the Challenging Collaboration of General Practitioners and Specialists for Patients with Complex Chronic Conditions: A Scoping Review. International Journal of Integrated Care, 22(3), 4. 10.5334/ijic.5970

Tricco, A. C., Zarin, W., Rios, P., Nincic, V., Khan, P. A., Ghassemi, M., Diaz, S., Pham, B., Straus, S. E., & Langlois, E. V. (2018). Engaging policy-makers, health system managers, and policy analysts in the knowledge synthesis process: a scoping review. Implementation Science, 13(1), 31. 10.1186/s13012-018-0717-x

Turner, J. P., Martin, P., Zhang, Y. Z., & Tannenbaum, C. (2020). Patients beliefs and attitudes towards deprescribing: Can deprescribing success be predicted? Research in Social and Administrative Pharmacy, 16(4), 599–604. 10.1016/j.sapharm.2019.07.007

Turner, J. P., Richard, C., Lussier, M.-T., Lavoie, M.-E., Farrell, B., Roberge, D., & Tannenbaum, C. (2018). Deprescribing conversations: a closer look at prescriber–patient communication. Therapeutic Advances in Drug Safety, 9(12), 687–698. 10.1177/2042098618804490

van Poelgeest, E. P., Seppala, L. J., Lee, J. M., Bahat, G., Ilhan, B., Lavan, A. H., Mair, A., van Marum, R. J., Onder, G., Ryg, J., Fernandes, M. A., Garfinkel, D., Guðmundsson, A., Hartikainen, S., Kotsani, M., Montero-Errasquín, B., Neumann-Podczaska, A., Pazan, F., Petrovic, M., … van der Velde, N. (2022). Deprescribing practices, habits and attitudes of geriatricians and geriatricians-in-training across Europe: a large web-based survey. European Geriatric Medicine, 13(6), 1455–1466. 10.1007/s41999-022-00702-9

Warkentin, L. M., Tjosvold, L., & Bond, K. (2026). An inventory of policy levers to reduce low value care: Results of a rapid scoping review. Health Policy, 164, 105508. 10.1016/j.healthpol.2025.105508

Wilkinson, H., Whittington, R., Perry, L., & Eames, C. (2017). Examining the relationship between burnout and empathy in healthcare professionals: A systematic review. Burnout Research, 6, 18–29. 10.1016/j.burn.2017.06.003

Zeynep Or, & Coralie Gandré. (2021). Sustainability and Resilience in the French Health System - World Economic Forum.

Zeynep Or, Coralie Gandré, Anna-Veera Seppänen, Cristina Hernández-Quevedo, Erin Webb, Morgane Michel, & Karine Chevreul. (2023). Health Systems in Transition. France - Health system review.

